# Identifying Potential Drug Targets for Varicose Veins through Integration of GWAS and eQTL Summary Data

**DOI:** 10.1101/2024.01.27.24301809

**Authors:** Yu Cui, Mengting Hu, He Zhou, Jiarui Guo, Qijia Wang, Zaihua Xu, Liyun Chen, Wancong Zhang, Shijie Tang

## Abstract

**Background:** Varicose veins (VV) is a common chronic venous disease that is influenced by multiple factors. It affects the quality of life of patients and imposes a huge economic burden on the healthcare system. This study aimed to use integrated analysis methods, including Mendelian randomization analysis, to identify potential pathogenic genes and drug targets for VV treatment.

**Methods:** This study conducted summary-data-based Mendelian randomization (SMR) analysis and colocalization analysis on data collected from genome-wide association studies (GWASs) and cis-expression quantitative trait loci (cis-eQTL) databases. Only genes with PP. H4 > 0.7 were chosen from the significant SMR results. After the above analysis, we screened 12 genes and performed Mendelian Randomization analysis on them. After sensitivity analysis, we identified four genes with potential causal relationships with VV. Finally, we used transcriptome-wide association studies (TWAS) analysis and The Drug-Gene Interaction Database (DGIdb, https://dgidb.org)data to identify and screen the remaining genes and identified four drug targets for the treatment of VV.

**Results:** We identified four genes significantly associated with VV, namely *KRTAP5-AS1* (OR = 1.08, 95% CI: 1.05−1.11, p = 1.42e−10) and *PLEKHA5* (OR = 1.13, 95% CI: 1.06-1.20, p = 6.90e−5), *CBWD1* (OR = 1.05, 95% CI: 1.01-1.11, p = 1.42e−2) and *CRIM1* (OR = 0.87, 95% CI: 0.81-0.95, p = 3.67e−3). Increased expression of three genes, namely *KRTAP5-AS1*, *PLEKHA5*, and *CBWD1*, was associated with increased risk of the disease, and increased expression of *CRIM1* was associated with decreased risk of the disease. These four genes could be targeted for VV therapy.

**Conclusion:** This study identified four drug targets for potential treatment of VV, and the results may contribute to improving the quality of life of VV patients.

## Introduction

Varicose veins (VV) are a common chronic venous disease affecting approximately one-third of the global population [1]. VV can manifest with a wide range of clinical presentations, ranging from asymptomatic to severe symptoms, including edema, pigmentation changes, eczema, lipodermatosclerosis, atrophie blanche, and healing or active venous ulcers [2]. The economic burden of VV on healthcare systems is substantial[3]. Current treatment approaches primarily involve pharmacotherapy, which is not always universally effective, necessitating more targeted therapeutic methods [4].

The incidence of VV is influenced by various factors, such as genetic susceptibility, environmental factors, hormones, endothelial dysfunction, activation of inflammatory cells and molecules, and disruption of cytokines and matrix metalloproteinase balance [5]. Observational epidemiological studies are prone to confounding, reverse causation, and various biases, which limit a deep understanding of the disease’s pathogenesis and treatment targets. However, Mendelian randomization (MR) methods, which are based on the random distribution of genetic polymorphisms, simulate the effects of randomized controlled trials and can eliminate the influence of confounding factors. In the context of VV research, this implies a more accurate assessment of the causal relationship between the drug targets and disease while excluding factors that could interfere with the results [6].

MR employs genetic variants as instrumental variables to estimate the causal effect of an exposure on the outcomes. It has been widely applied in other disease studies and helped successfully identify therapeutic targets for various diseases [7,8]. However, there is limited research on employing MR methods to explore potential drug targets for VV. Therefore, to gain a deeper understanding of the pathogenesis of VV and identify more effective treatment approaches, further MR studies are needed to evaluate the drug targets for VV. This will help eliminate factors that could interfere with the results and offer new perspectives and methods for the treatment of VV.

Conceptually, single nucleotide polymorphisms (SNPs) are randomly distributed and not influenced by environmental factors, making them an ideal tool for causal inference. MR is a form of instrumental variable analysis that primarily utilizes SNPs as genetic instruments to estimate the causal effect of an exposure (in this case, circulatory proteins) on the outcomes [9]. MR has been successfully used in previous studies to identify biomarkers and treatment targets for various diseases, including aortic aneurysms [10], multiple sclerosis [11], and breast cancer [12].

In the MR analysis of drug targets, cis-expression quantitative trait loci (cis-eQTLs) located in the drug target gene regions are often considered proxies that act as regulators of gene expression. Previous research has identified potential pathogenic proteins for VV, such as *IRF3*, *LUM*, *POSTN*, *RSPO3*, and *SARS2*, through a drug target MR analysis that focused on 2,004 plasma proteins [13]. However, this study had significant limitations, and the analysis of drug targets for VV remains insufficient, with numerous potential drug targets awaiting discovery. To address this research gap, our study focuses on conducting drug-target research with genes as the exposure factor for VV. Unlike previous studies that used proteins as the exposure factor, we explored the role of genes in the development of VV and as potential drug targets. Research using genes as the exposure factor has several advantages. First, genes have stable genetic characteristics, and they are unaffected by environmental factors, allowing for a more accurate assessment of the causal relationship between genes and VV [14]. Second, using genes as the exposure factor can provide more direct information and help reveal potential pathogenic mechanisms and drug targets for VV [15]. By exploring VV’s potential drug targets through genes as the exposure factor, we aim to offer new perspectives and methods for the treatment and prevention of VV.

In this study, our goal was to identify potential pathogenic genes for VV. We conducted summary-data-based Mendelian randomization (SMR), MR, and transcriptome-wide association study (TWAS) analyses by combining eQTLs identified in blood with independent genome-wide association study (GWAS) datasets for varicose veins.

## Materials and methods

**Fig. 1.**
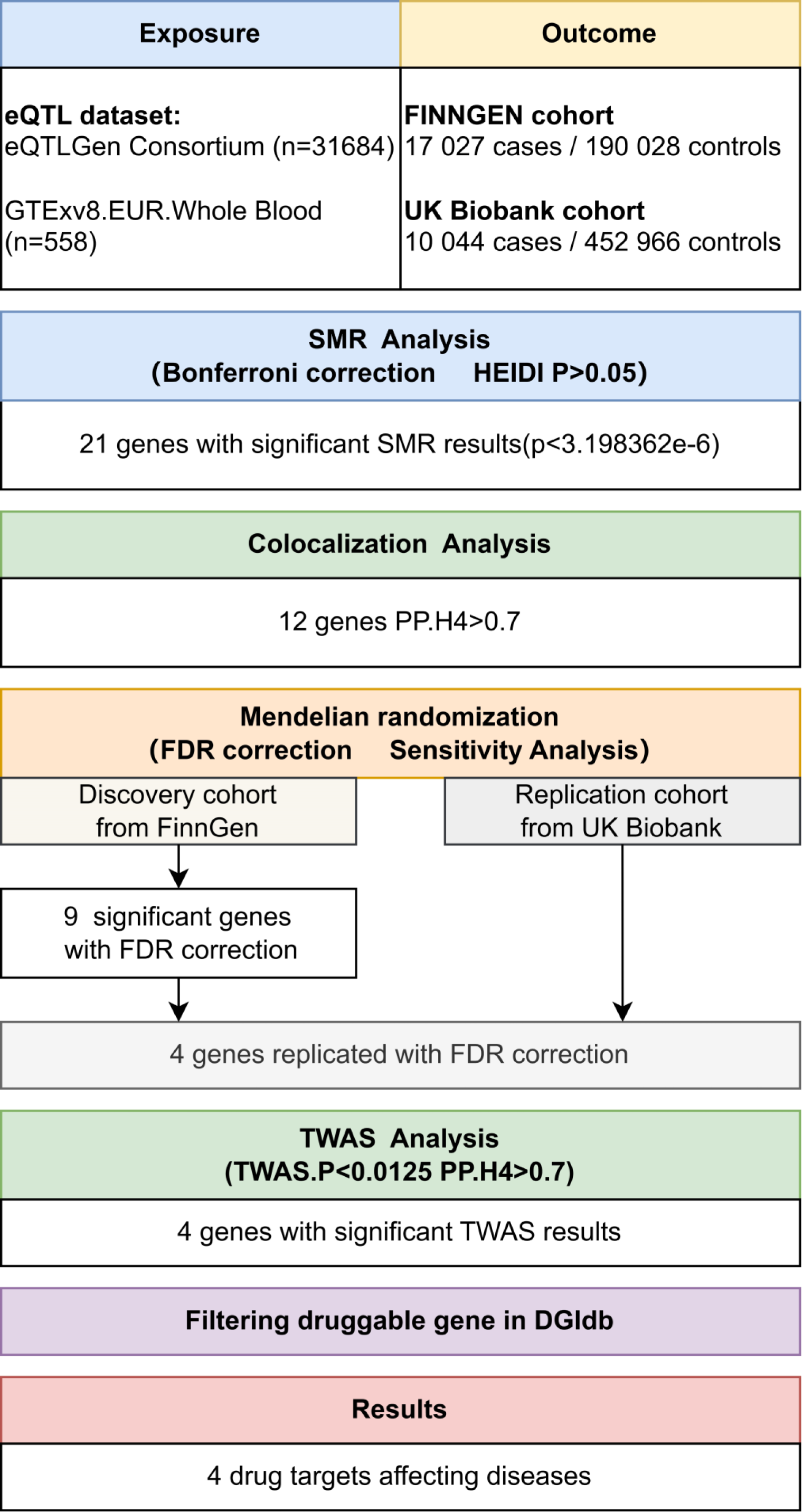
Overview of the study design.

### Datasets

#### GWAS

We obtained summary-level GWAS data on VV from the FinnGen consortium, which comprise 17,027 cases with VV and 190,028 controls. To the best of our knowledge, these data constitute the most recent GWAS findings, encompassing the largest number of VV cases to date. The overarching objective of FinnGen is to accumulate and meticulously analyze the genome and national health register data from 500,000 Finnish individuals [16].

For external replication with an independent sample cohort, we used the summary statistics from the UK Biobank. The UK Biobank is a large-scale biomedical database that encompasses genetic and health information of more than 500,000 participants in the United Kingdom. This resource provides a valuable platform for researchers and offers extensive data on various health conditions, lifestyle factors, and genetic profiles. In our study, the UK Biobank dataset included 10,044 patients with VV and 452,966 controls and contributed to the robustness and generalizability of our findings [17,18].

#### eQTL

In our study’s context of drug development, we focused on cis-eQTLs that were in proximity to the target gene. These cis-eQTLs for the SMR analysis were sourced from the eQTLGen Consortium and the eQTL meta-analysis conducted on peripheral blood samples from a cohort of 31,684 individuals [19]. These selected cis-eQTLs served as valuable inputs for our SMR, colocalization [20], and MR analyses.

For the TWAS analysis of the eQTL data, we used the GTEx v8 European whole blood dataset. GTEx is a notable biomedical research endeavor dedicated to comprehending the diversity in human gene expression across diverse tissues and organs to elucidate its intricate connections with genotype. Given our specific emphasis on VV, we meticulously extracted comprehensive eQTL results exclusively from the whole blood samples within the GTEx dataset [21].

#### DGIdb

The Drug Gene Interaction Database (DGIdb, www.DGIdb.org) is as a comprehensive drug-gene interaction network resource. It amalgamates various data sources that elucidate the interactions between drugs and genes [22] as well as the pharmaceutical relevance of genes. In this study, we used the DGIdb database for the identification of candidate gene targets for gene therapy from the previously identified genes.

#### SMR analysis

We conducted summary data-based Mendelian randomization (SMR) and heterogeneity in dependent instruments (HEIDI) tests analyses on cis regions using the SMR software (version 1.03). The methodologies for SMR analysis are detailed in the original work by Z et al. [23]. In brief, SMR analysis uses a well-established MR approach. This technique employs a SNP at a prominent cis-eQTL as an instrumental variable (IV). The summary-level eQTL data act as the exposure, and the GWAS data for a specific trait serve as the outcome. The primary objective is to explore a potential causal or pleiotropic association, wherein the same causal variant is shared between gene expression and the trait.

It is crucial to acknowledge that the SMR method lacks the ability to distinguish between a causal association, in which gene expression causally influences the trait, and a pleiotropic association, in which the same SNP affects both gene expression and the trait. This limitation arises due to the single IV in the MR method, which cannot differentiate causality from pleiotropy. Nevertheless, the HEIDI test can make this distinction by discerning causality and pleiotropy from linkage. Linkage denotes cases in which two different SNPs in linkage disequilibrium (LD) independently impact gene expression and the trait. Although less biologically intriguing than causality and pleiotropy, the HEIDI test brings clarity in such scenarios.

For the SMR tests, statistical significance was set at a p value less than 3.198362 × 10^−6 (Bonferroni correction of 0.05 divided by 15,633, the number of results in the eQTL data). For the HEIDI test, a p value below 0.05 was considered significant and suggesting that the observed association was attributable to linkage [23].

#### Colocalization analysis

We conducted colocalization analysis using the coloc package within the R software environment (version 4.0.3) [20]. The purpose of colocalization analysis is to evaluate whether SNPs associated with both gene expression and phenotype at a given locus are indeed shared causal variants, indicating the “colocalization” of gene expression and the phenotype. The analysis, implemented through colocalization analysis, computes posterior probabilities (PPs) for five hypotheses: 1) H0, indicating no association with either gene expression or the phenotype; 2) H1, denoting an association solely with gene expression; 3) H2, signifying an association exclusively with the phenotype; 4) H3, suggesting an association with both gene expression and the phenotype through independent SNPs; and 5) H4, implying an association with both the gene expression and phenotype through shared causal SNPs. A substantial PP for H4 (PP.H4 above 0.70) strongly indicates the existence of shared causal variants influencing both gene expression and the phenotype [20].

#### MR analysis

To perform the two-sample MR analysis [18], we used the TwoSampleMR R package (version 0.5.6, accessible at https://mrcieu.github.io/TwoSampleMR/). The application of the two-sample MR framework requires the use of two distinct datasets. In this study, genetic instruments, specifically cis-eQTL, were employed as the exposures, and GWAS data served as the outcome trait data. The MR methodology evaluates the association between gene expression and diseases or traits by leveraging genetic variants linked to gene expression as the instrumental variables (exposure) and GWAS as the outcomes. MR allows for the exploration of whether changes in gene expression causally influence diseases or traits. For instruments represented by a SNP, the Wald ratio was applied. In the cases in which the instruments comprised multiple SNPs, the inverse variance-weighted MR approach was implemented.

The MR analysis was exclusively conducted for the results of the SMR and colocalization analyses. We applied the false discovery rate (FDR) method to correct the p values. The analysis of pleiotropy employed the MR_pleiotropy_test function, where a p value less than 0.05 was considered to indicate the presence of pleiotropy, and a p value greater than 0.05 indicated its absence. Significance parameters were set to p < 5 × 10^−8 for genome-wide significance; the linkage disequilibrium parameter (r^2) was set to 0.1, and the genetic distance was set to 10 Mb.

#### TWAS analysis

TWAS is a research approach employed to establish connections between the gene expression levels and intricate networks governing gene expression. By scrutinizing the extensive transcriptomic datasets, researchers can discern distinct gene expression patterns across varied biological conditions. This method enables the identification of pivotal genes linked to specific physiological processes or diseases while elucidating intricate gene interactions. Through these investigations, essential genes crucial to distinct biological processes are pinpointed, revealing their regulatory networks and potential signaling pathways [24].

In this study, a TWAS fusion analysis was conducted on autosomal chromosomes while adhering to the default parameters outlined in the TWAS fusion analysis guidelines (http://gusevlab.org/projects/fusion/). Colocalization assessment was performed using the coloc R package for the genes that had transcriptome-wide significance and resided within a 1.5 Mb range. This Bayesian methodology computes posterior probabilities (PPs), which signifies the likelihood of shared causal variants within a locus for two outcomes. This analytical approach distinguishes associations influenced by horizontal pleiotropy (a solitary causal SNP impacting both transcription and VV; posterior probability PP4) from those arising due to linkage (two causal SNPs in LD affecting transcription and VV independently; posterior probability PP3) [20].

## Results

### SMR analysis and colocalization for Preliminary Identifying Potential Genes

In the initial phase of our analysis, we conducted SMR analysis using the eQTLGen dataset to identify genes significantly associated with VV features. To establish the significance threshold for the p values, we applied the Bonferroni method to establish the threshold for the *p* values. Specifically, for the eQTLGen dataset, we obtained a wealth of information, resulting in 15,633 candidate genes. To maintain a robust level of significance, we set the significance threshold at (0.05 / 15,633 = 3.198362e−06). In the SMR analysis, we identified 21 genes (*SLC2A1-AS1, CRIM1, PDK1, AC007401.2, AC093818.1, HMCES, HSPA4, TRIM10, HCG22, NOS3, CBWD1, ZNF438, ARHGEF17, ADM, KRTAP5-AS1, PLEKHA5, NFATC3, PPL, DPEP3, MAP2K4,* and *LSM4*) that met the significance threshold of p < 3.198362 E-06. To enhance interpretability, Manhattan plots (refer to Fig. 2) were generated for a clear visualization of the SMR results.

**Fig. 2.**
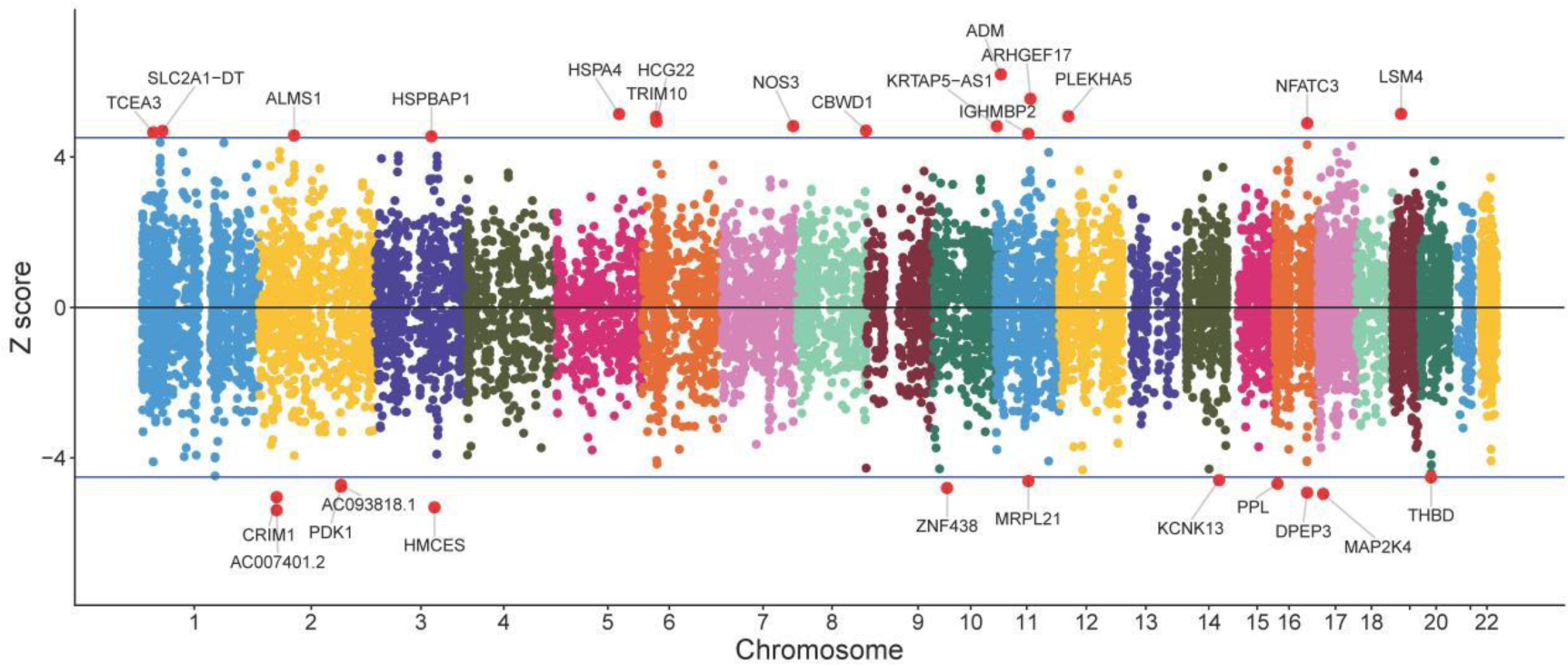
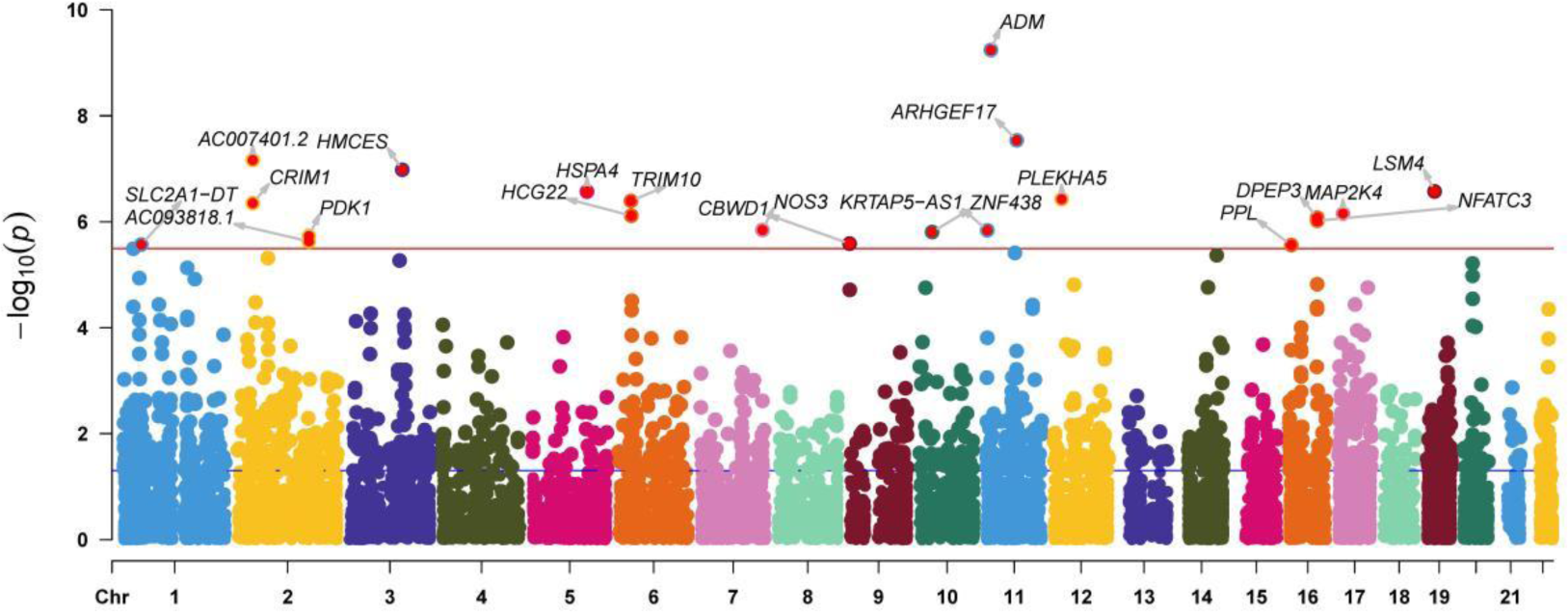
Manhattan plot of the SMR analysis results using QTLs and VV GWAS summary statistics. The red dashed line represents the Bonferroni-corrected significance threshold (Sig_P_Thresh). Z-score filtration was applied, with the blue dashed line indicating the Z score significance level, where Sig_Z_Thresh was calculated as qnorm (1 - [0.05 / Sig_P_Thresh]).

**Fig. 3.**
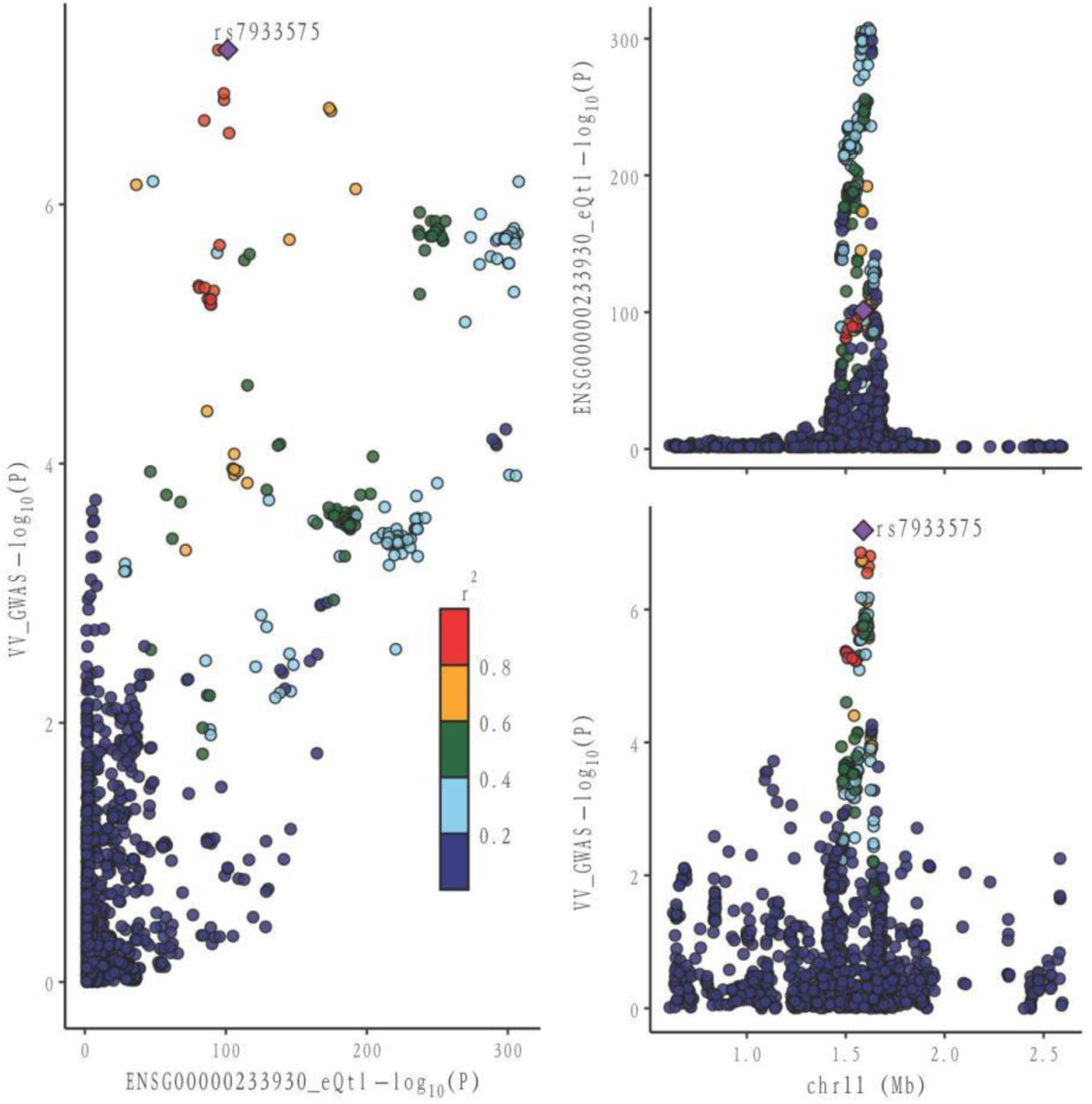
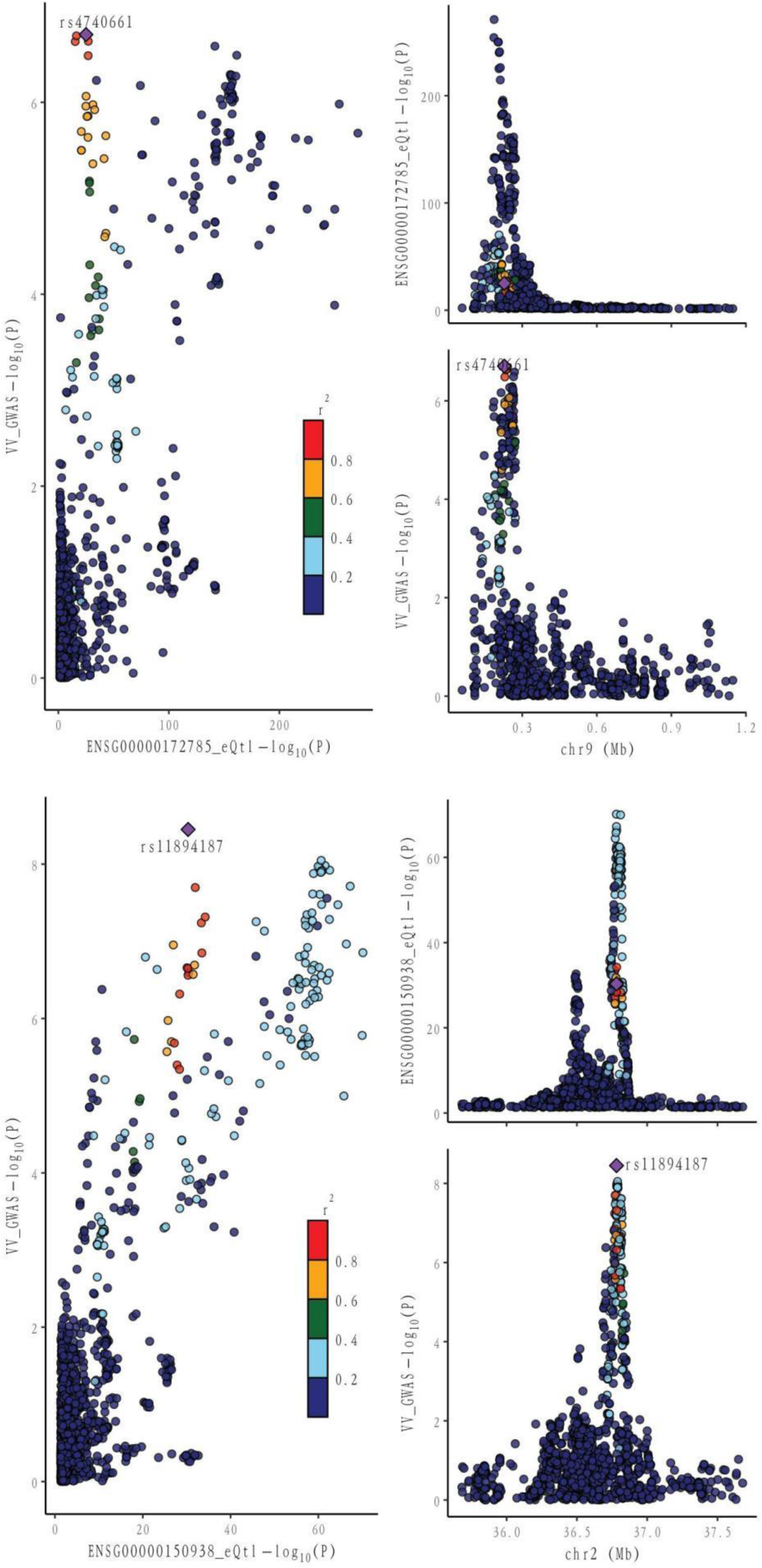
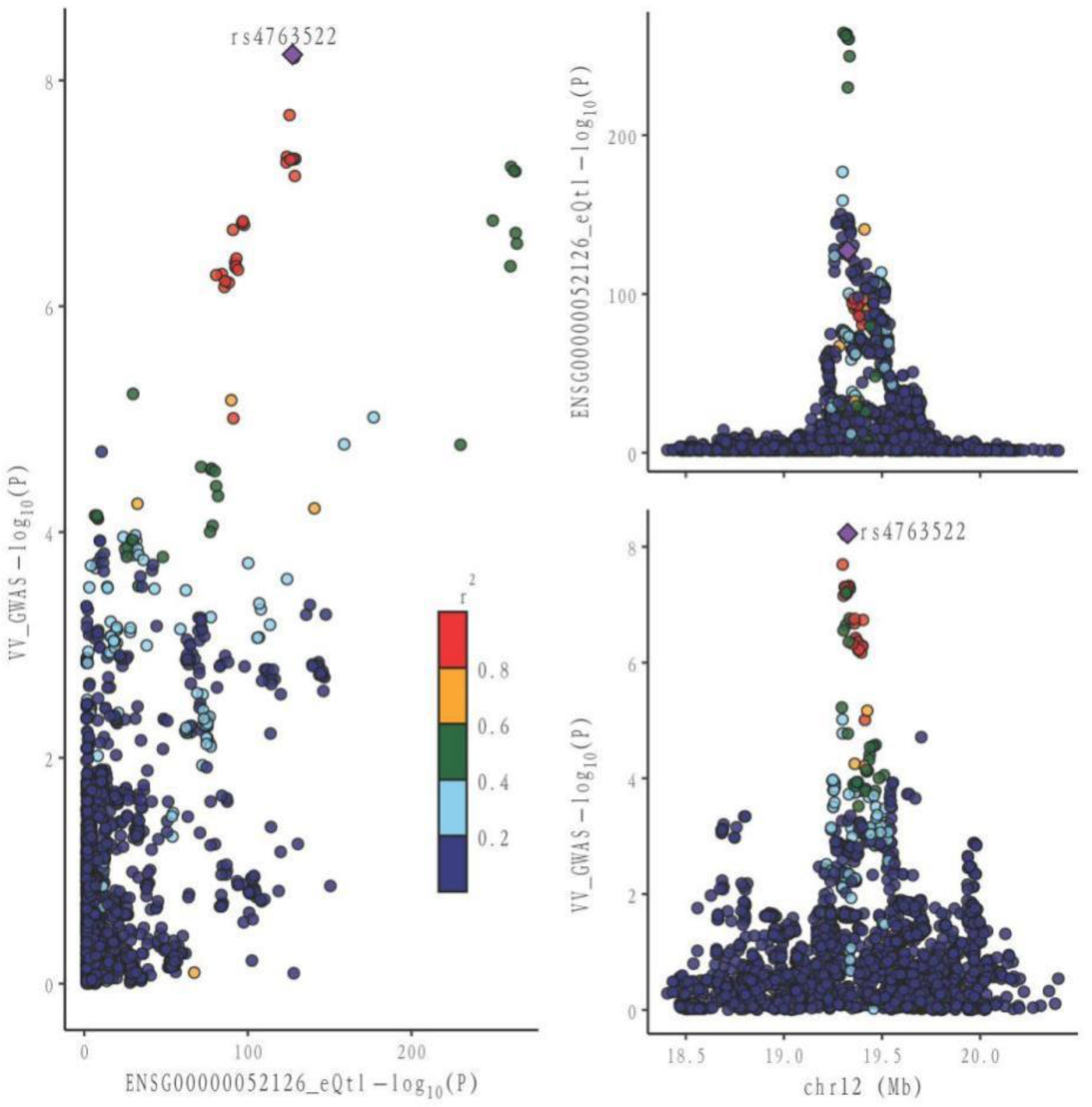
Locus comparison plot illustrating the colocalization analysis results for the single nucleotide variants associated with gene expression in both blood and VV data. Each dot on the plot represents a SNP, and its color corresponds to the LD value (r2) with the GWAS lead variant, depicted as a purple diamond. In the right panel, the genomic positions (in megabases, GRCh37) on the chromosome are presented on the x-axis. The −log10 p values for SNPs from the VV GWAS (top) and the eQTL study for the genes (bottom) are displayed on the y-axis. In the left panel, a comparison of p values is shown for both the VV GWAS and the gene expression eQTL study.

Following the identification of these significant genes, we employed the HEIDI heterogeneity test (p_HEIDI > 0.05) to filter out genes lacking horizontal pleiotropy. In summary, through the integrated analysis of GWAS and blood eQTL data, we identified a total of 21 genes significantly associated with VV features in blood (refer to Table 1).

**Table 1.**
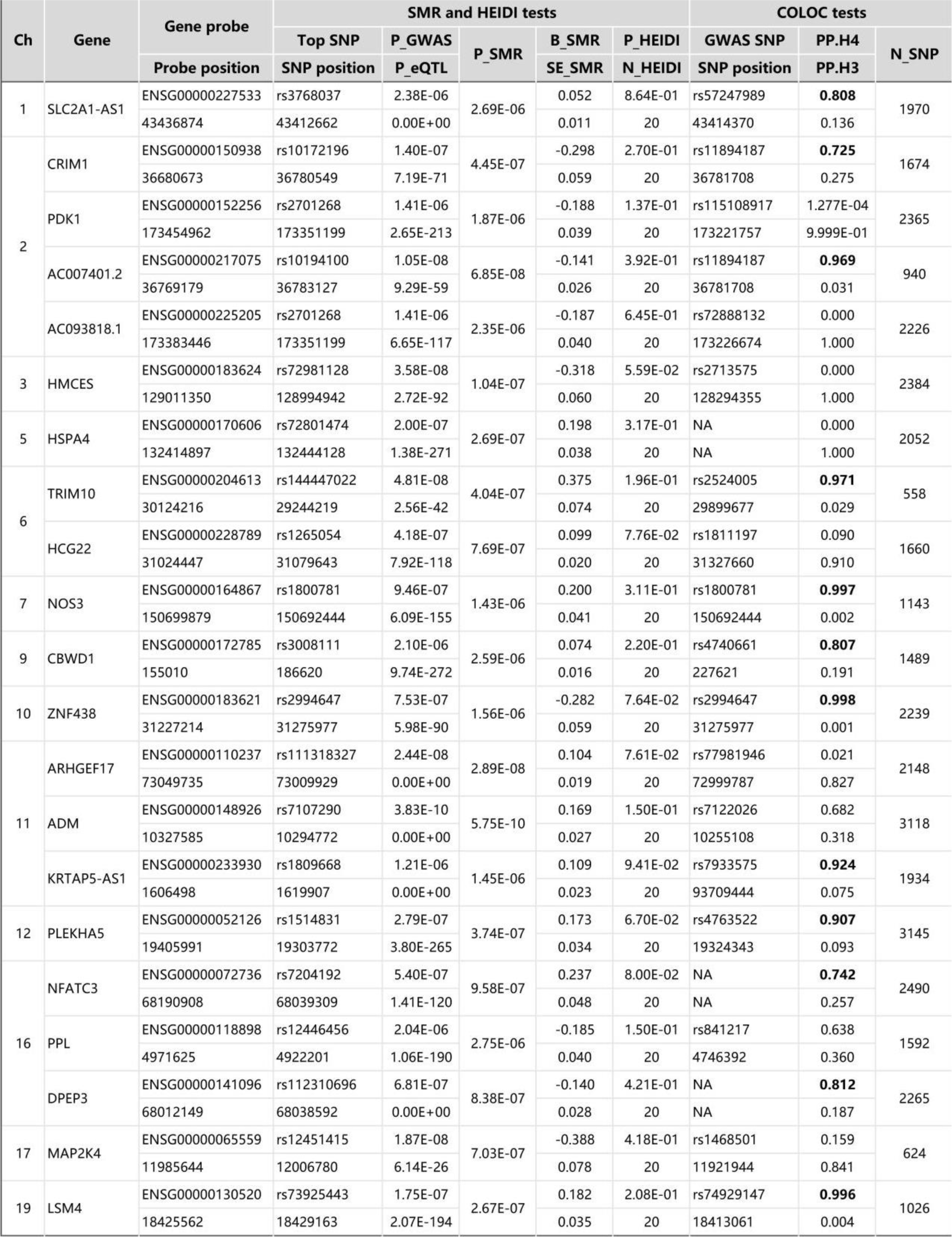
SMR/HEIDI results of the GWAS data on VV, blood eQTL data, and colocalization results between GWAS and the blood eQTL data for genes passing the SMR Test. Probe and SNP positions are indicated in GRCh37. Bold numbers denote large PP.H4 (> 0.70). Ch represents chromosome; PGWAS is the p-value of the top SNP from the GWAS data; P_eQTL is the p-value of the top SNP from the eQTL data; P_SMR is the p-value for the SMR test; B_SMR is the effect size from the SMR test; SE_SMR is the standard error of B_SMR; P_HEIDI is the p-value for the HEIDI test; N_HEIDL is the number of SNPs used in the HEIDI test; GWAS SNP is the lead variant with the smallest p-value from the GWAS data in the region analyzed by the colocalization test (±1 Mb from the GWAS SNP position); N_SNP is the number of SNPs used in the colocalization test.

Moving forward, we conducted colocalization analysis to integrate the GWAS and blood eQTL data for the genes that successfully passed the SMR test. This analysis aimed to assess whether these genes were colocalized with the trait of interest, VV. The results from the colocalization test provided robust evidence supporting colocalization between the trait and all 12 genes (*KRTAP5-AS1*, *SLC2A1-DT*, *PLEKHA5*, *NOS3*, *LSM4*, *DPEP3*, *CRIM1*, *ZNG1A*, *ZNF438*, *NFATC3*, *TRIM10*, and *AC007401.2*) that met the criteria of both the SMR and HEIDI tests (Table 1, results with bold numbers in PP.H4). Therefore, we consider these genes as high-priority candidates for subsequent functional studies.

### MR analysis Validates Potential Gene Causal Relationships

#### Discovery Cohort

Utilizing cis-eQTL data provided by the eQTLGen Consortium and by employing SMR and colocalization analyses, we identified 12 potential genes that could influence VV. Subsequently, a two-sample MR analysis was conducted on the European summary statistics involving patients with VV. In the discovery cohort, which comprised 17,027 patients and 190,028 controls from the FinnGen cohort, the inverse variance weighted (IVW) MR analysis amalgamated the effect estimates from each genetic instrument. The genetically predicted expression of nine genes was found to be associated with VV risk, after multiple testing adjustments (FDR correction). In order to ensure the robustness of our findings, we performed tests for horizontal pleiotropy, which indicated no evidence of horizontal pleiotropy in the dataset.

In our study, we performed a two-sample MR analysis using the 12 genes. The results are presented in Table 2. In the discovery cohort, significant associations were observed for nine genes (*KRTAP5-AS1, SLC2A1-DT, NOS3, PLEKHA5, LSM4, DPEP3, CRIM1, AC007401.2,* and *ZNG1A*). Heterogeneity tests and horizontal pleiotropy tests were conducted. The heterogeneity test revealed heterogeneity for *PLEKHA5* and *CBWD1*. Consequently, we applied the random effects inverse variance weighted method to address this heterogeneity in the subsequent analysis. The results of the horizontal pleiotropy analysis indicated the absence of horizontal pleiotropy. These additional details provide a nuanced understanding of the robustness and validity of our findings in the context of VV genetics.

**Table 2.**
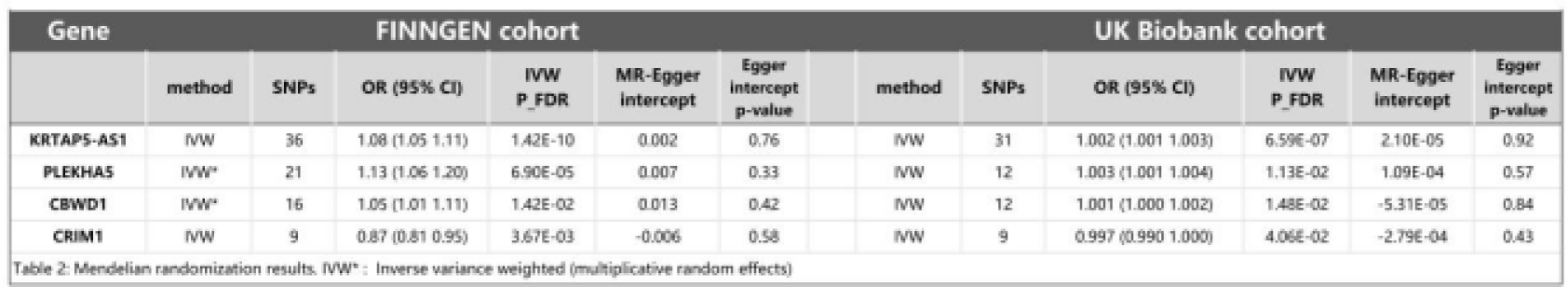
MR results.

**Table 3.**
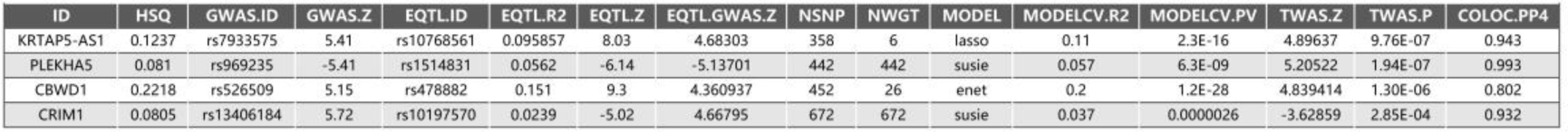
TWAS results between GWAS and blood eQTL data for genes through the TWAS/Colocalization Test. HSQ stands for gene heritability. GWAS.ID and GWAS.Z are the top GWAS SNPs rsIDs and Z-scores. EQTL.ID and EQTL.R2 are the best eQTLs rsIDs and cross-validated r^2^. NSNP is the locus SNP count. MODEL is the best-performing model. TWAS.Z and TWAS.P are key TWAS statistics: Z score and p value. COLOC.PP4 represents the colocalization analysis results.

#### Replication Cohort

In our endeavor to validate the genetic associations identified in the discovery phase, we conducted a rigorous replication analysis using data from the UK Biobank cohort, comprising 10,044 cases and 452,966 controls. This replication effort aimed to reproduce the effect estimates of the top nine genes identified in the discovery stage, providing crucial insights into the robustness and generalizability of our findings.

We observed significant associations with VV for *KRTAP5-AS1* (Odds ratio (OR) = 1.08, 95% CI: 1.05-1.11, p = 1.42e−10), *PLEKHA5* (OR = 1.13, 95% CI: 1.06-1.20, p = 6.90e-5), *CBWD1* (OR = 1.05, 95% CI: 1.01-1.11, p = 1.42e-2), and *CRIM1* (OR = 0.87, 95% CI: 0.81-0.95, p = 3.67e-3). Notably, these findings underscore the robustness and consistency of the genetic associations across different cohorts, reinforcing the potential relevance of these genes to VV.

Of particular significance, our replication analysis demonstrated successful replication beyond the false discovery rate (FDR) method for four genes (*KRTAP5-AS1, PLEKHA5, CBWD1,* and *CRIM1*), exhibiting 100% consistency in the direction of the effect. This remarkable consistency further validates the genetic associations and supports the reliability of our findings. Importantly, horizontal pleiotropy tests indicated no evidence suggesting the presence of this confounding factor in the data, further bolstering the robustness and validity of our replicated genetic associations with VV.

#### TWAS analysis Validates Transcriptome-level Causal Relationships

In our pursuit of strengthening the causal inference and gaining deeper insights into the genetic associations with VV, we conducted a Transcriptome-Wide Association Study (TWAS) on the four genes identified in the previous analyses. This comprehensive approach aimed to elucidate the transcriptional level associations of these genes with VV and to bolster the evidence supporting their potential causal role. Additionally, we performed colocalization analysis to further validate the potential causal relationship between these genes and VV.

The results revealed significant associations at the transcriptional level for the four genes with VV, with all colocalization probabilities (PP.H4.p) exceeding 0.7. Specifically, elevated transcription levels of *KRTAP5-AS1* (Z score = 4.896, p = 9.76e−7), *PLEKHA5* (Z score = 5.205, p = 1.94e−7), and *CBWD1* (Z score = 4.839, p = 1.30e−6) were significantly correlated with an increased risk of VV, whereas increased transcription level of *CRIM1* (Z score = −3.629, p = 2.85e−4) was significantly associated with a decreased risk of VV. Notably, these TWAS findings were consistent with those from the SMR analysis, providing robust support for the transcriptional associations of these genes with VV.

We also conducted a TWAS colocalization analysis and found that the probability of these four genes sharing the same causal variant (PP.H4.p: *KRTAP5-AS1* = 0.943, *PLEKHA5* = 0.993, *CBWD1* = 0.802, *CRIM1* = 0.932) exceeded the predefined threshold (p > 0.7).This robust result unequivocally strengthens the indication of the causal relationship between these four genes and VV, further solidifying the genetic basis of VV and providing crucial insights into the potential mechanisms underlying this condition.

#### DGIdb Screening for Pharmacologically Relevant Genes

Incorporating the comprehensive resources provided by the DGIdb database, our analysis identified four pharmacologically relevant genes (*KRTAP5-AS1, PLEKHA5, CBWD1,* and *CRIM1*). This crucial step has unveiled potential drug targets with direct relevance to the treatment of VV, shedding light on promising avenues for targeted therapeutic interventions. By leveraging this pharmacogenomic information, we have advanced our understanding of the molecular underpinnings of VV and have laid the groundwork for the exploration of novel therapeutic strategies tailored to the genetic landscape of this condition.

## Discussion

To study the potential pathogenic genes of VV and to search for new drug targets, we used an integrated analysis method combining SMR analysis and colocalization analysis. We used an eQTL meta-analysis dataset based on peripheral blood samples and a DNA methylation quantitative trait loci (mQTLs) dataset as the exposure data with a relatively large sample size for the outcome data from two large databases, FinnGen and UK Biobank. After the analysis of the preliminary results, we screened out some candidate genes and identified them using the DGIdb database. Finally, four VV-related genes were identified: *KRTAP5-AS1*, *PLEKHA5, CBWD1*, and *CRIM1*. The results suggested that there was a potential causal relationship between VV and these four genes. The increased expression of *KRTAP5-AS1, PLEKHA5,* and *CBWD1* was associated with increased risk of VV, and the increased expression of *CRIM1* was associated with a significantly decreased risk of VV. The results of this study may provide important evidence for drug development and individualized treatment of VV and contribute to improving the quality of life of VV patients.

There are few studies on *KRTAP5-AS1* and *CBWD1*. The research on *KRTAP5-AS1* is mainly focused on its long-chain non-coding RNA (lncRNA), which is a kind of conserved RNA with a length of more than 200 nucleotides and no significant protein-coding ability. There is evidence that it can be used as a new biomarker to predict the prognosis of various cancers [25]. For example, Song et al. found that *KRTAP5-AS1* can exert the role of a cRNA in regulating claudin-4, and that *KRTAP5-AS1* can regulate *CLDN4* expression as a competitive endogenous RNA of miR-596 and miR-3620-3p. *CLDN4* has been shown to alter expression patterns in various types of cancer, including gastric, pancreatic, and ovarian cancer [26]. In addition, studies have used *KRTAP5-AS1* as a lncRNA signal to predict the prognosis of papillary thyroid cancer [25]. Other studies have found the potential of eight lncRNA markers, including *KRTAP5-AS1*, as independent prognostic biomarkers in HBV-positive hepatocellular carcinoma patients [27].

*CBWD1* is located at 9p24.3:121038-179075 (GRCH37 / HG19) and located near the 9p telomere. The absence of 9p syndrome can lead to facial deformity, hypotonia, and mental retardation. Researchers have found that *CBWD1* is associated with some cases of congenital kidney and urinary tract abnormalities. The results have suggested that *CBWD1* is associated with the development of ureteric buds into the urinary tract, and homozygous deletion involving *CBWD1* can manifest as renal hypoplasia [28]. Similar to *KRTAP5-AS1*, there is an association between *CBWD1* and cancer, as reported by Wang et al., who constructed a brisk tmmodel of *CBWD1* as a pivotal gene to predict survival in patients with ovarian cancer [29]. Most existing studies on *KRTAP5-AS1* and *CBWD1* have been in the field of cancer, and there is no evidence of a potential link between these two genes and VV. However, our results suggest that this could be a research direction. *PLEKHA5* and *CRIM1* are two relatively popular genes that have been studied extensively. *PLEKHA5* belongs to the *PLEKHA* family and contains the pleckstring homology domain. The domain is thought to mediate phosphatidylinositol binding properties and is associated with multiple intracellular functions, such as signaling, cytoskeleton rearrangement, membrane protein targeting, and vesicle trafficking [30]. *PLEKHA5* has been shown to be expressed on the plasma membrane and associated with microtubules, and it has also been demonstrated to play an important role in cell migration by wound healing assay experiments [31]. To date, the most thoroughly studied aspect of *PLEKHA5* has been its potential regulatory role in melanoma brain metastasis. *PLEKHA5* is associated with brain cell activity, and decreased expression of *PLEKHA5* inhibited the proliferation of brain nutrient cells as well as the migration of pro-brain cells in an in vitro blood-brain barrier model. *PLEKHA5* expression in melanoma is associated with the early development of brain metastases, and up to 75% of patients with stage IV melanoma develop central nervous system metastases during the course of the disease, according to a study by Jilaveanu et al [32]. *PLEKHA5* has also been reported to be involved in the central nervous system homing mechanism in metastatic disease, and inhibition of *PLEKHA5* may reduce the passage across the blood-brain barrier and reduce the proliferation and survival of melanoma cells in the brain and extrabrain regions [32]. Since there is no experimental evidence to support this, we can only speculate that *PLEKHA5*’s positive causal relationship to VV may also be mediated by brain metastasis. In addition to melanoma, *PLEKHA5* has also been associated with gastric cancer, with studies finding that tyrosine phosphorylation of *PLEKHA5* is *MET*-dependent and associated with *MET* expression and phosphorylation. *PLEKHA5* is able to regulate the survival and peritoneal dissemination of diffuse gastric cancer cells through *MET* gene amplification [33].

*CRIM1* is a gene encoding a cysteine-rich repeat protein that is developmentally regulated and involved in vertebrate central nervous system development and organogenesis [34]. Evidence that *CRIM1* haploinsufficiency leads to defects in eye development in humans and mice suggests *CRIM1* as a potential causative gene of Macrophthalmia, colobomatous, with microcornea (MACOM) and underscores the importance of *CRIM1* in eye development [35]. In addition, *CRIM1* is also associated with the formation and maintenance of blood vessels in vivo. During angiogenesis, *CRIM1* is strongly upregulated in endothelial cells and expressed by a variety of cell lines with adhesion growth, and the formation of capillary structures is impaired in endothelial cells that are transfected in vitro [36]. We hypothesize that the increased expression of *CRIM1* might reduce the risk of VV because the endothelial structure of blood vessels is not susceptible to damage.

In the existing research on *PLEKHA5* and *CRIM1*, we did not find any studies linking *PLEKHA5* and *CRIM1* with VV. Our experimental results demonstrate the existence of a potential causal relationship between *PLEKHA5* and *CRIM1* and VV, but the possible pathways or mechanisms are unknown.

In the past, dissection of the great saphenous vein and the small saphenous vein, which often occur in VV, were the main treatment. However, this method was risky, and the recurrence rate was relatively high, and up to 24% of patients required additional treatment. As a result, minimally invasive procedures such as radiofrequency ablation, intravenous laser therapy, ultrasound-guided foam sclerosis, and others started to be favored [37]. Minimally invasive surgery has also been shown to be relatively safe with few serious complications [38]. Our results are at the genetic level, and if we can understand the pathways or mechanisms by which these genes function, we may be able to introduce a new type of therapy based on new bioscience techniques.

There are some limitations in this study. First, our study was only related to theoretical drug target discovery. Second, the two-sample MR analysis used in the study was based on publicly available and free GWAS aggregates with limitations in updating, selection, and screening of the data. In addition, although we used a variety of methods to validate the results of our experiments, we cannot rule out the possibility that other confounding factors that are not taken into account may have influenced the effects of the genes on the disease. Further exploration and validation is required.

In conclusion, this study identified the effects of *KRTAP5-AS1, PLEKHA5, CBWD1,* and *CRIM1* genes on VV. No previous study has found a potential causal relationship between these four genes and VV. Our results provide new causal evidence for VV-related genes as well as potential drug targets and therapeutic ideas for VV therapy.

## Data Availability

All data produced are available online from each genome-wide association study.

https://gwas.mrcieu.ac.uk/datasets/ukb-b-6720/

https://yanglab.westlake.edu.cn/data/SMR/GTEx_V8_cis_eqtl_summary.html

https://www.eqtlgen.org/cis-eqtls.html

https://storage.googleapis.com/finngen-public-data-r9/summary_stats/finngen_R9_I9_VARICVE.gz

## Acknowledgements

Not applicable

## References

[1] R. de Ávila Oliveira, R. Riera, V. Vasconcelos, J.C. Baptista-Silva, Injection sclerotherapy for varicose veins, Cochrane Database Syst. Rev. 12 (2021) CD001732. 10.1002/14651858.CD001732.pub3.

[2] Y.J. Youn, J. Lee, Chronic venous insufficiency and varicose veins of the lower extremities, Korean J. Intern. Med. 34 (2019) 269–283. 10.3904/kjim.2018.230.

[3] R.D. Malgor, N. Labropoulos, Diagnosis and follow-up of varicose veins with duplex ultrasound: how and why?, Phlebology 27 Suppl 1 (2012) 10–15. 10.1258/phleb.2011.012s05.

[4] A. Belramman, R. Bootun, T.R.A. Lane, A.H. Davies, Foam sclerotherapy versus ambulatory phlebectomy for the treatment of varicose vein tributaries: study protocol for a randomised controlled trial, Trials 20 (2019) 392. 10.1186/s13063-019-3398-0.

[5] J.D. Raffetto, Pathophysiology of Chronic Venous Disease and Venous Ulcers, Surg. Clin. North Am. 98 (2018) 337–347. 10.1016/j.suc.2017.11.002.

[6] G. Davey Smith, G. Hemani, Mendelian randomization: genetic anchors for causal inference in epidemiological studies, Hum. Mol. Genet. 23 (2014) R89–98. 10.1093/hmg/ddu328.

[7] M.K. Georgakis, D. Gill, Mendelian Randomization Studies in Stroke: Exploration of Risk Factors and Drug Targets With Human Genetic Data, Stroke 52 (2021) 2992–3003. 10.1161/STROKEAHA.120.032617.

[8] C. Y, Y. Y, H. Q, W. G, Identification of potential drug targets for rheumatoid arthritis from genetic insights: a Mendelian randomization study, J. Transl. Med. 21 (2023). 10.1186/s12967-023-04474-z.

[9] B.-B. M, E. Us, Z. X, B. Ao, A. Hj, R. Aa, C. A, C. We, M. R, N. K, F. S, R. A, W. L, L. E, P.F. None, G. S, B. H, H. Im, T. Me, N. G, Z. Mc, High-coverage whole-genome sequencing of the expanded 1000 Genomes Project cohort including 602 trios, Cell 185 (2022). 10.1016/j.cell.2022.08.004.

[10] Y. Chen, X. Xu, L. Wang, K. Li, Y. Sun, L. Xiao, J. Dai, M. Huang, Y. Wang, D.W. Wang, Genetic insights into therapeutic targets for aortic aneurysms: A Mendelian randomization study, eBioMedicine 83 (2022) 104199. 10.1016/j.ebiom.2022.104199.

[11] J. Lin, J. Zhou, Y. Xu, Potential drug targets for multiple sclerosis identified through Mendelian randomization analysis, Brain 146 (2023) 3364–3372. 10.1093/brain/awad070.

[12] Y. Wang, F. Liu, L. Sun, Y. Jia, P. Yang, D. Guo, M. Shi, A. Wang, G.-C. Chen, Y. Zhang, Z. Zhu, Association between human blood metabolome and the risk of breast cancer, Breast Cancer Res. 25 (2023) 9. 10.1186/s13058-023-01609-4.

[13] J. Lin, J. Zhou, Z. Liu, R. Zeng, L. Wang, F. Li, L. Cui, Y. Zheng, Identification of potential drug targets for varicose veins: a Mendelian randomization analysis, Front. Cardiovasc. Med. 10 (2023) 1126208. 10.3389/fcvm.2023.1126208.

[14] S. Shah, A. Henry, C. Roselli, H. Lin, G. Sveinbjörnsson, G. Fatemifar, Å.K. Hedman, J.B. Wilk, M.P. Morley, M.D. Chaffin, A. Helgadottir, N. Verweij, A. Dehghan, P. Almgren, C. Andersson, K.G. Aragam, J. Ärnlöv, J.D. Backman, M.L. Biggs, H.L. Bloom, J. Brandimarto, M.R. Brown, L. Buckbinder, D.J. Carey, D.I. Chasman, X. Chen, X. Chen, J. Chung, W. Chutkow, J.P. Cook, G.E. Delgado, S. Denaxas, A.S. Doney, M. Dörr, S.C. Dudley, M.E. Dunn, G. Engström, T. Esko, S.B. Felix, C. Finan, I. Ford, M. Ghanbari, S. Ghasemi, V. Giedraitis, F. Giulianini, J.S. Gottdiener, S. Gross, D.F. Guðbjartsson, R. Gutmann, C.M. Haggerty, P. van der Harst, C.L. Hyde, E. Ingelsson, J.W. Jukema, M. Kavousi, K.-T. Khaw, M.E. Kleber, L. Køber, A. Koekemoer, C. Langenberg, L. Lind, C.M. Lindgren, B. London, L.A. Lotta, R.C. Lovering, J. Luan, P. Magnusson, A. Mahajan, K.B. Margulies, W. März, O. Melander, I.R. Mordi, T. Morgan, A.D. Morris, A.P. Morris, A.C. Morrison, M.W. Nagle, C.P. Nelson, A. Niessner, T. Niiranen, M.L. O’Donoghue, A.T. Owens, C.N.A. Palmer, H.M. Parry, M. Perola, E. Portilla-Fernandez, B.M. Psaty, Regeneron Genetics Center, K.M. Rice, P.M. Ridker, S.P.R. Romaine, J.I. Rotter, P. Salo, V. Salomaa, J. van Setten, A.A. Shalaby, D.T. Smelser, N.L. Smith, S. Stender, D.J. Stott, P. Svensson, M.-L. Tammesoo, K.D. Taylor, M. Teder-Laving, A. Teumer, G. Thorgeirsson, U. Thorsteinsdottir, C. Torp-Pedersen, S. Trompet, B. Tyl, A.G. Uitterlinden, A. Veluchamy, U. Völker, A.A. Voors, X. Wang, N.J. Wareham, D. Waterworth, P.E. Weeke, R. Weiss, K.L. Wiggins, H. Xing, L.M. Yerges-Armstrong, B. Yu, F. Zannad, J.H. Zhao, H. Hemingway, N.J. Samani, J.J.V. McMurray, J. Yang, P.M. Visscher, C. Newton-Cheh, A. Malarstig, H. Holm, S.A. Lubitz, N. Sattar, M.V. Holmes, T.P. Cappola, F.W. Asselbergs, A.D. Hingorani, K. Kuchenbaecker, P.T. Ellinor, C.C. Lang, K. Stefansson, J.G. Smith, R.S. Vasan, D.I. Swerdlow, R.T. Lumbers, Genome-wide association and Mendelian randomisation analysis provide insights into the pathogenesis of heart failure, Nat. Commun. 11 (2020) 163. 10.1038/s41467-019-13690-5.

[15] M. Crous-Bou, I. De Vivo, C.A. Camargo, R. Varraso, F. Grodstein, M.K. Jensen, P. Kraft, S.Z. Goldhaber, S. Lindström, C. Kabrhel, Interactions of established risk factors and a GWAS-based genetic risk score on the risk of venous thromboembolism, Thromb. Haemost. 116 (2016) 705–713. 10.1160/TH16-02-0172.

[16] M.I. Kurki, J. Karjalainen, P. Palta, T.P. Sipilä, K. Kristiansson, K.M. Donner, M.P. Reeve, H. Laivuori, M. Aavikko, M.A. Kaunisto, A. Loukola, E. Lahtela, H. Mattsson, P. Laiho, P. Della Briotta Parolo, A.A. Lehisto, M. Kanai, N. Mars, J. Rämö, T. Kiiskinen, H.O. Heyne, K. Veerapen, S. Rüeger, S. Lemmelä, W. Zhou, S. Ruotsalainen, K. Pärn, T. Hiekkalinna, S. Koskelainen, T. Paajanen, V. Llorens, J. Gracia-Tabuenca, H. Siirtola, K. Reis, A.G. Elnahas, B. Sun, C.N. Foley, K. Aalto-Setälä, K. Alasoo, M. Arvas, K. Auro, S. Biswas, A. Bizaki-Vallaskangas, O. Carpen, C.-Y. Chen, O.A. Dada, Z. Ding, M.G. Ehm, K. Eklund, M. Färkkilä, H. Finucane, A. Ganna, A. Ghazal, R.R. Graham, E.M. Green, A. Hakanen, M. Hautalahti, Å.K. Hedman, M. Hiltunen, R. Hinttala, I. Hovatta, X. Hu, A. Huertas-Vazquez, L. Huilaja, J. Hunkapiller, H. Jacob, J.-N. Jensen, H. Joensuu, S. John, V. Julkunen, M. Jung, J. Junttila, K. Kaarniranta, M. Kähönen, R. Kajanne, L. Kallio, R. Kälviäinen, J. Kaprio, FinnGen, N. Kerimov, J. Kettunen, E. Kilpeläinen, T. Kilpi, K. Klinger, V.-M. Kosma, T. Kuopio, V. Kurra, T. Laisk, J. Laukkanen, N. Lawless, A. Liu, S. Longerich, R. Mägi, J. Mäkelä, A. Mäkitie, A. Malarstig, A. Mannermaa, J. Maranville, A. Matakidou, T. Meretoja, S.V. Mozaffari, M.E.K. Niemi, M. Niemi, T. Niiranen, C.J. O Donnell, M.E. Obeidat, G. Okafo, H.M. Ollila, A. Palomäki, T. Palotie, J. Partanen, D.S. Paul, M. Pelkonen, R.K. Pendergrass, S. Petrovski, A. Pitkäranta, A. Platt, D. Pulford, E. Punkka, P. Pussinen, N. Raghavan, F. Rahimov, D. Rajpal, N.A. Renaud, B. Riley-Gillis, R. Rodosthenous, E. Saarentaus, A. Salminen, E. Salminen, V. Salomaa, J. Schleutker, R. Serpi, H.-Y. Shen, R. Siegel, K. Silander, S. Siltanen, S. Soini, H. Soininen, J.H. Sul, I. Tachmazidou, K. Tasanen, P. Tienari, S. Toppila-Salmi, T. Tukiainen, T. Tuomi, J.A. Turunen, J.C. Ulirsch, F. Vaura, P. Virolainen, J. Waring, D. Waterworth, R. Yang, M. Nelis, A. Reigo, A. Metspalu, L. Milani, T. Esko, C. Fox, A.S. Havulinna, M. Perola, S. Ripatti, A. Jalanko, T. Laitinen, T.P. Mäkelä, R. Plenge, M. McCarthy, H. Runz, M.J. Daly, A. Palotie, FinnGen provides genetic insights from a well-phenotyped isolated population, Nature 613 (2023) 508–518. 10.1038/s41586-022-05473-8.

[17] A. Fry, T.J. Littlejohns, C. Sudlow, N. Doherty, L. Adamska, T. Sprosen, R. Collins, N.E. Allen, Comparison of Sociodemographic and Health-Related Characteristics of UK Biobank Participants With Those of the General Population, Am. J. Epidemiol. 186 (2017) 1026–1034. 10.1093/aje/kwx246.

[18] G. Hemani, J. Zheng, B. Elsworth, K.H. Wade, V. Haberland, D. Baird, C. Laurin, S. Burgess, J. Bowden, R. Langdon, V.Y. Tan, J. Yarmolinsky, H.A. Shihab, N.J. Timpson, D.M. Evans, C. Relton, R.M. Martin, G. Davey Smith, T.R. Gaunt, P.C. Haycock, The MR-Base platform supports systematic causal inference across the human phenome, eLife 7 (2018) e34408. 10.7554/eLife.34408.

[19] U. Võsa, A. Claringbould, H.-J. Westra, M.J. Bonder, P. Deelen, B. Zeng, H. Kirsten, A. Saha, R. Kreuzhuber, S. Yazar, H. Brugge, R. Oelen, D.H. de Vries, M.G.P. van der Wijst, S. Kasela, N. Pervjakova, I. Alves, M.-J. Favé, M. Agbessi, M.W. Christiansen, R. Jansen, I. Seppälä, L. Tong, A. Teumer, K. Schramm, G. Hemani, J. Verlouw, H. Yaghootkar, R. Sönmez Flitman, A. Brown, V. Kukushkina, A. Kalnapenkis, S. Rüeger, E. Porcu, J. Kronberg, J. Kettunen, B. Lee, F. Zhang, T. Qi, J.A. Hernandez, W. Arindrarto, F. Beutner, BIOS Consortium, i2QTL Consortium, J. Dmitrieva, M. Elansary, B.P. Fairfax, M. Georges, B.T. Heijmans, A.W. Hewitt, M. Kähönen, Y. Kim, J.C. Knight, P. Kovacs, K. Krohn, S. Li, M. Loeffler, U.M. Marigorta, H. Mei, Y. Momozawa, M. Müller-Nurasyid, M. Nauck, M.G. Nivard, B.W.J.H. Penninx, J.K. Pritchard, O.T. Raitakari, O. Rotzschke, E.P. Slagboom, C.D.A. Stehouwer, M. Stumvoll, P. Sullivan, P.A.C. ’t Hoen, J. Thiery, A. Tönjes, J. van Dongen, M. van Iterson, J.H. Veldink, U. Völker, R. Warmerdam, C. Wijmenga, M. Swertz, A. Andiappan, G.W. Montgomery, S. Ripatti, M. Perola, Z. Kutalik, E. Dermitzakis, S. Bergmann, T. Frayling, J. van Meurs, H. Prokisch, H. Ahsan, B.L. Pierce, T. Lehtimäki, D.I. Boomsma, B.M. Psaty, S.A. Gharib, P. Awadalla, L. Milani, W.H. Ouwehand, K. Downes, O. Stegle, A. Battle, P.M. Visscher, J. Yang, M. Scholz, J. Powell, G. Gibson, T. Esko, L. Franke, Large-scale cis- and trans-eQTL analyses identify thousands of genetic loci and polygenic scores that regulate blood gene expression, Nat. Genet. 53 (2021) 1300–1310. 10.1038/s41588-021-00913-z.

[20] C. Giambartolomei, D. Vukcevic, E.E. Schadt, L. Franke, A.D. Hingorani, C. Wallace, V. Plagnol, Bayesian test for colocalisation between pairs of genetic association studies using summary statistics, PLoS Genet. 10 (2014) e1004383. 10.1371/journal.pgen.1004383.

[21] GTEx Consortium, The GTEx Consortium atlas of genetic regulatory effects across human tissues, Science 369 (2020) 1318–1330. 10.1126/science.aaz1776.

[22] A.H. Wagner, A.C. Coffman, B.J. Ainscough, N.C. Spies, Z.L. Skidmore, K.M. Campbell, K. Krysiak, D. Pan, J.F. McMichael, J.M. Eldred, J.R. Walker, R.K. Wilson, E.R. Mardis, M. Griffith, O.L. Griffith, DGIdb 2.0: mining clinically relevant drug-gene interactions, Nucleic Acids Res. 44 (2016) D1036–1044. 10.1093/nar/gkv1165.

[23] Z. Z, Z. F, H. H, B. A, R. Mr, P. Je, M. Gw, G. Me, W. Nr, V. Pm, Y. J, Integration of summary data from GWAS and eQTL studies predicts complex trait gene targets, Nat. Genet. 48 (2016). 10.1038/ng.3538.

[24] A. Gusev, A. Ko, H. Shi, G. Bhatia, W. Chung, B.W.J.H. Penninx, R. Jansen, E.J.C. de Geus, D.I. Boomsma, F.A. Wright, P.F. Sullivan, E. Nikkola, M. Alvarez, M. Civelek, A.J. Lusis, T. Lehtimäki, E. Raitoharju, M. Kähönen, I. Seppälä, O.T. Raitakari, J. Kuusisto, M. Laakso, A.L. Price, P. Pajukanta, B. Pasaniuc, Integrative approaches for large-scale transcriptome-wide association studies, Nat. Genet. 48 (2016) 245–252. 10.1038/ng.3506.

[25] X. You, S. Yang, J. Sui, W. Wu, T. Liu, S. Xu, Y. Cheng, X. Kong, G. Liang, Y. Yao, Molecular characterization of papillary thyroid carcinoma: a potential three-lncRNA prognostic signature, Cancer Manag. Res. 10 (2018) 4297–4310. 10.2147/CMAR.S174874.

[26] Y.-X. Song, J.-X. Sun, J.-H. Zhao, Y.-C. Yang, J.-X. Shi, Z.-H. Wu, X.-W. Chen, P. Gao, Z.-F. Miao, Z.-N. Wang, Non-coding RNAs participate in the regulatory network of CLDN4 via ceRNA mediated miRNA evasion, Nat. Commun. 8 (2017) 289. 10.1038/s41467-017-00304-1.

[27] X. Zhao, Z. Bai, C. Li, C. Sheng, H. Li, Identification of a Novel Eight-lncRNA Prognostic Signature for HBV-HCC and Analysis of Their Functions Based on Coexpression and ceRNA Networks, BioMed Res. Int. 2020 (2020) 8765461. 10.1155/2020/8765461.

[28] S. Kanda, M. Ohmuraya, H. Akagawa, S. Horita, Y. Yoshida, N. Kaneko, N. Sugawara, K. Ishizuka, K. Miura, Y. Harita, T. Yamamoto, A. Oka, K. Araki, T. Furukawa, M. Hattori, Deletion in the Cobalamin Synthetase W Domain-Containing Protein 1 Gene Is associated with Congenital Anomalies of the Kidney and Urinary Tract, J. Am. Soc. Nephrol. JASN 31 (2020) 139–147. 10.1681/ASN.2019040398.

[29] H. Wang, J. Liu, J. Yang, Z. Wang, Z. Zhang, J. Peng, Y. Wang, L. Hong, A novel tumor mutational burden-based risk model predicts prognosis and correlates with immune infiltration in ovarian cancer, Front. Immunol. 13 (2022) 943389. 10.3389/fimmu.2022.943389.

[30] S. Dowler, R.A. Currie, D.G. Campbell, M. Deak, G. Kular, C.P. Downes, D.R. Alessi, Identification of pleckstrin-homology-domain-containing proteins with novel phosphoinositide-binding specificities, Biochem. J. 351 (2000) 19–31. 10.1042/0264-6021:3510019.

[31] Y. Zou, W. Zhong, A likely role for a novel PH-domain containing protein, PEPP2, in connecting membrane and cytoskeleton, Biocell Off. J. Soc. Latinoam. Microsc. Electron. Al 36 (2012) 127–132.

[32] L.B. Jilaveanu, F. Parisi, M.L. Barr, C.R. Zito, W. Cruz-Munoz, R.S. Kerbel, D.L. Rimm, M.W. Bosenberg, R. Halaban, Y. Kluger, H.M. Kluger, PLEKHA5 as a Biomarker and Potential Mediator of Melanoma Brain Metastasis, Clin. Cancer Res. Off. J. Am. Assoc. Cancer Res. 21 (2015) 2138–2147. 10.1158/1078-0432.CCR-14-0861.

[33] Y. Nagamura, M. Miyazaki, Y. Nagano, M. Yuki, K. Fukami, K. Yanagihara, K. Sasaki, R. Sakai, H. Yamaguchi, PLEKHA5 regulates the survival and peritoneal dissemination of diffuse-type gastric carcinoma cells with Met gene amplification, Oncogenesis 10 (2021) 25. 10.1038/s41389-021-00314-1.

[34] G. Kolle, K. Georgas, G.P. Holmes, M.H. Little, T. Yamada, CRIM1, a novel gene encoding a cysteine-rich repeat protein, is developmentally regulated and implicated in vertebrate CNS development and organogenesis, Mech. Dev. 90 (2000) 181–193. 10.1016/s0925-4773(99)00248-8.

[35] F. Beleggia, Y. Li, J. Fan, N.H. Elcioğlu, E. Toker, T. Wieland, I.H. Maumenee, N.A. Akarsu, T. Meitinger, T.M. Strom, R. Lang, B. Wollnik, CRIM1 haploinsufficiency causes defects in eye development in human and mouse, Hum. Mol. Genet. 24 (2015) 2267–2273. 10.1093/hmg/ddu744.

[36] J. Glienke, A. Sturz, A. Menrad, K.H. Thierauch, CRIM1 is involved in endothelial cell capillary formation in vitro and is expressed in blood vessels in vivo, Mech. Dev. 119 (2002) 165–175. 10.1016/s0925-4773(02)00355-6.

[37] T. Nijsten, R.R. van den Bos, M.P. Goldman, M.A. Kockaert, T.M. Proebstle, E. Rabe, N.S. Sadick, R.A. Weiss, M.H.A. Neumann, Minimally invasive techniques in the treatment of saphenous varicose veins, J. Am. Acad. Dermatol. 60 (2009) 110–119. 10.1016/j.jaad.2008.07.046.

[38] A. Al Samaraee, I.J.D. McCallum, A. Mudawi, Endovenous therapy of varicose veins: a better outcome than standard surgery?, Surg. J. R. Coll. Surg. Edinb. Irel. 7 (2009) 181–186. 10.1016/s1479-666x(09)80043-7.

